# CAPICE: a computational method for Consequence-Agnostic Pathogenicity Interpretation of Clinical Exome variations

**DOI:** 10.1101/19012229

**Authors:** Shuang Li, K. Joeri van der Velde, Dick de Ridder, Aalt D.J. van Dijk, Dimitrios Soudis, Leslie R. Zwerwer, Patrick Deelen, Dennis Hendriksen, Bart Charbon, Marielle van Gijn, Kristin M. Abbott, B. Sikkema-Raddatz, Cleo C. van Diemen, Wilhelmina S. Kerstjens-Frederikse, Richard J. Sinke, Morris A. Swertz

**Author notes:** Corresponding author: Morris Swertz. equal contribution (Shuang Li, K. Joeri van der Velde).

## Abstract

Exome sequencing is now mainstream in clinical practice, however, identification of pathogenic Mendelian variants remains time consuming, partly because limited accuracy of current computational prediction methods leaves much manual classification. Here we introduce CAPICE, a new machine-learning based method for prioritizing pathogenic variants, including SNVs and short InDels, that outperforms best general (CADD, GAVIN) and consequence-type-specific (REVEL, ClinPred) computational prediction methods, for both rare and ultra-rare variants. CAPICE is easily integrated into diagnostic pipelines and is available as free and open source command-line software, file of pre-computed scores, and as a web application with web service API.

## BACKGROUND

The past decades have seen rapid advances in genetic testing and increasing numbers of trial studies aimed at using genetic testing to facilitate rare disease diagnostics, and many studies have demonstrated the unique role whole exome and genome sequencing can play in improving diagnostic yield [(1), (2), (3), (4), (5), (6), (7)]. However, the vast amount of genomic data that is now available has created large interpretation challenges that can be alleviated using computational tools. However, variant interpretation in particular still remains time-consuming, in part because of the limited accuracy of current computational prediction methods and the manual work required to identify large numbers of false positives produced by those methods [(8), (9), (10)].

Existing prediction methods can be categorized into two groups. One group of methods [(11), (12)] focuses on specific types of variants, with the majority of these methods only classifying non-synonymous single nucleotide variants (nsSNVs) [(13), (14)]. Successful methods of this group include Clinpred (15), which has the best current performance validated in multiple datasets, and REVEL [(16)], which specifically targets rare variants. However, these methods miss the diagnosis when the causal variant is not an nsSNV, which is the case for 76% of reported pathogenic variants (17). The other category of prediction methods provides predictions of selective constraints for a broader range of variations [(18), (19), (20), (21)] that can also inform pathogenicity classification. A method that is widely used and acknowledged for performance is CADD [(22)], which estimates the deleteriousness of SNVs and short insertions and deletions (InDels). However, these tools are built for estimating evolutionary constraints and do not directly target pathogenicity. They can also introduce ascertainment bias for variants that are under high evolutionary pressure (such as nonsense and splicing variants) even though these can be observed in healthy populations, and they can neglect rare and recent variants that have not undergone purifying selection but are still found to contribute to diseases [(23)].

New computational prediction methods need to be examined for their ability to reduce the number of variants that requires time-consuming expert evaluation as this is currently a bottleneck in the diagnostic pipeline. With hundreds to thousands of non-pathogenic variants identified in a typical patient with a rare genetic disorder, it is important to restrict the false positive rate of computational prediction methods, i.e. the number of neutral variants falsely reported as pathogenic. However, new methods are currently often not evaluated for their ability to recognize neutral variants. Indeed, a recent review (24) found that commonly used variant interpretation tools may incorrectly predict a third of the common variations found in the Exome Aggregation Consortium (ExAC) to be harmful. We speculate that this may be explained by the bias in training data selection because the neutral set used in different tools can be biased towards common neutral variants [(15), (25), (26)], which in practice means that the pathogenicity of rare and ultra-rare variants cannot be accurately estimated. Therefore, it is important to avoid bias in data selection and evaluate false positive rate of the prediction methods in clinical setting where rare and ultra-rare neutral variants are frequently encountered using neutral benchmark datasets [(27), (28)] and clinical data.

The challenge for rare disease research and diagnostics is thus to find robust classification algorithms that perform well for all the different types of variants and allele frequencies. To meet this challenge, we developed CAPICE, a new method for <u>C</u>onsequence-<u>A</u>gnostic prediction of <u>P</u>athogenicity Interpretation of <u>C</u>linical <u>E</u>xome variations. CAPICE overcomes limitations common in current predictors by training a sophisticated machine learning model that targets (non-)pathogenicity, using a specifically prepared, high confidence and pathogenicity versus benign balanced training dataset, and using many existing genomic annotations across the entire genome (the same features that were used to produce CADD). In high quality benchmark sets CAPICE thus outperforms existing methods in distinguishing pathogenic variants from neutral variants, irrespective of their different molecular consequences and allele frequency and, to our knowledge, CAPICE is the first and only variant prioritization method that targets pathogenicity prediction of all-types of SNVs and InDels, irrespective of consequence type.

Below we will describe the results of our performance evaluations, discuss features and limitations of our methodology, provide extensive details on the materials and methods used, concluding that CAPICE thus offers high accuracy pathogenicity classification across all consequence types and allele frequencies, outperforming all next-best variant classification methods. To make CAPICE easy to access, we have developed CAPICE as both a command-line tool and a web-app, and released it with pre-computed scores available as ready-to-use annotation files.

## RESULTS

Below we report performance analysis of CAPICE compared to the best current prediction models using gold standard benchmark sets, analysis of the classification consistency of CAPICE across different allele frequency ranges and across different types of variants and a small practical evaluation where we applied CAPICE to a set of patient exomes.

### CAPICE outperforms the best current prediction methods

CAPICE is a general prediction method that provides pathogenicity estimations for SNVs and InDels across different molecular consequences (Figure *1*). In our performance comparison, we included recently published prediction methods and those that show best performance in benchmark studies. In case a tool was not able to provide a prediction we marked it as ‘No prediction returned’. Because most prediction methods are built specifically for non-synonymous variants, we performed the comparison for both the full dataset and the non-synonymous subset. In our benchmark datasets, CAPICE performs as well or better than other current prediction methods across all categories (*Figure 1, Supplementary Figure 3, Supplementary Figure 4, Supplementary Table 1, Supplementary Table 2*). We also examined the robustness of CAPICE’s performance for rare and ultra-rare variants and variants that lead to different consequences.

**Figure 1:**
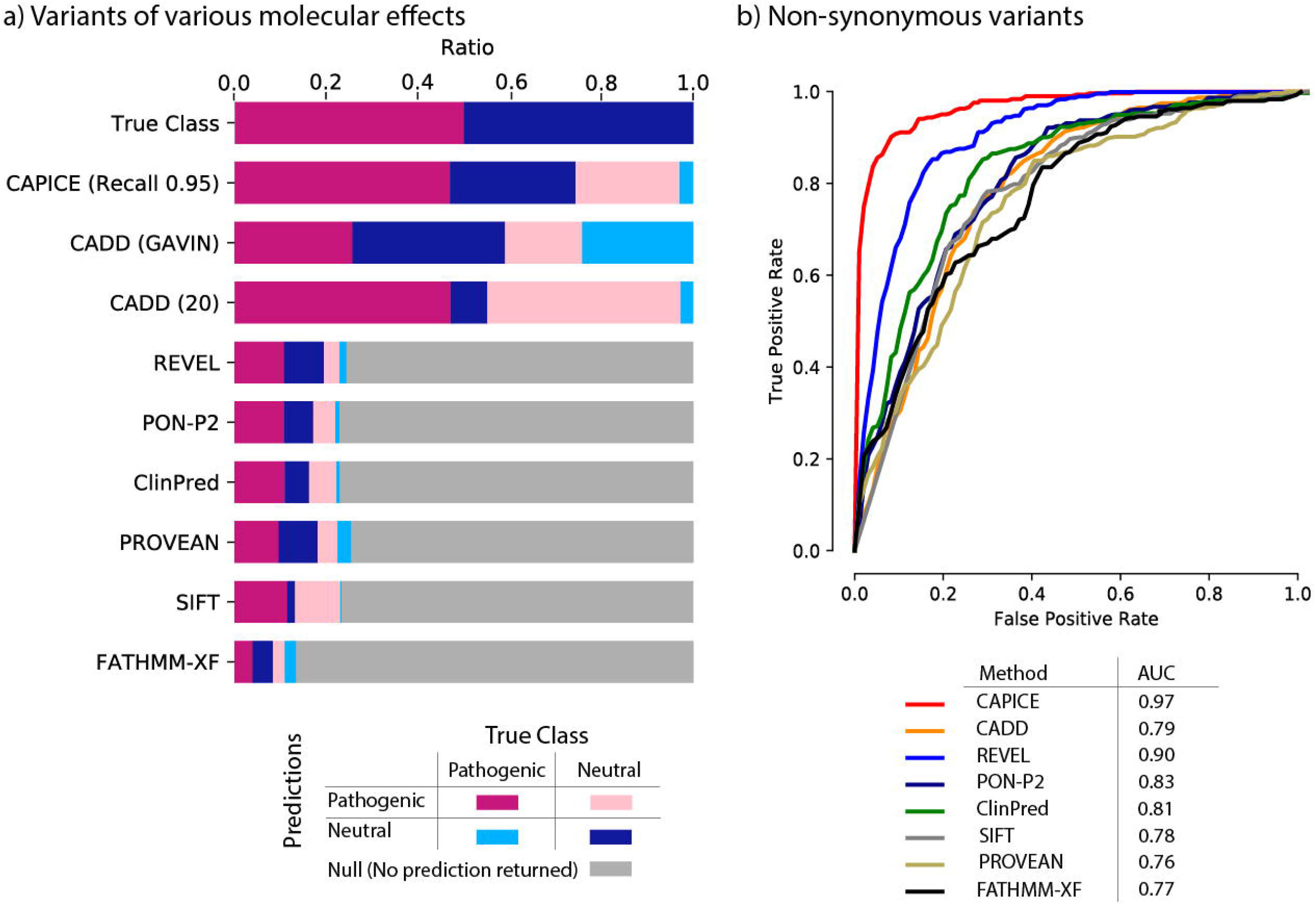
CAPICE outperforms other predictors in discriminating pathogenic variants and neutral variants. a) True/false classification for all predictors tested against the full benchmark set that contains all types of variants. Top bar shows the breakdown of the test set. Other bars show the classification performance for each method. Purple blocks represent correct classification of pathogenic variants. Dark-blue blocks represent neutral variants. Pink and light-blue blocks denote false classifications. Gray blocks represent variants that were not classified by the predictor tested. Threshold-selection methods are in Method section. b) Receiver operating characteristic (ROC) curves of CAPICE with AUC values for a subset of the benchmark data that only contains non-synonymous variants (ROC curve for the full dataset can be found in Supplementary Figure 3). Each ROC curve is for a subset of variants displaying a specific molecular consequence. AUC values for the different methods are listed in the figure legend.

For the full data, CAPICE outperformed CADD, the mostly used ‘general’ prediction method, and achieved an area under the receiver operating characteristic curve (AUC) of 0.89 as compared to 0.53 for CADD (shown in Supplementary Figure *3*). For the non-synonymous subset, CAPICE outperformed all the other prediction methods and achieved an AUC of 0.97 (shown in Figure *1b*). The majority of other methods we examined are built specifically for non-synonymous variants, with the exception of FATHMM-XF, which was developed for point mutations. For the non-synonymous subset, REVEL, which was built for rare variants, produced the second best result and achieved an AUC of 0.90.

To asses impact of these difference in practice we assumed a clinical setting with the aim to recognize 95% of the pathogenic variants (which is a very high standard in current practice). When using a threshold of 0.02 on CAPICE classification score, CAPICE correctly recognized 95% of pathogenic variants in the full test dataset and wrongly classified 50% of the neutral variants as pathogenic – which was the lowest number of misclassified variants among all the predictors we tested. In contrast, CADD with a score threshold of 20 achieved a comparable recall of 94%, but wrongly classified 85% of neutral variants as pathogenic. When using gene-specific CADD score thresholds based on the GAVIN method (29), the performance of CADD was better but still much worse than CAPICE. All other tested methods could give predictions less than 30% of the full dataset.

We also examined how well the prediction methods can recognize neutral variants in two neutral benchmark datasets. For both datasets, CAPICE’s performance was comparable to or better than the current best prediction methods (Supplementary Table *2*, Supplementary Table ***3***).

### CAPICE outperforms other current predictors for rare and ultra-rare variants

CAPICE performs consistently across different allele frequencies and especially well for rare and ultra-rare variants. Here we repeated the evaluation strategy for the same benchmark dataset grouped into five allele frequency bins (Figure *2*).

**Figure 2.**
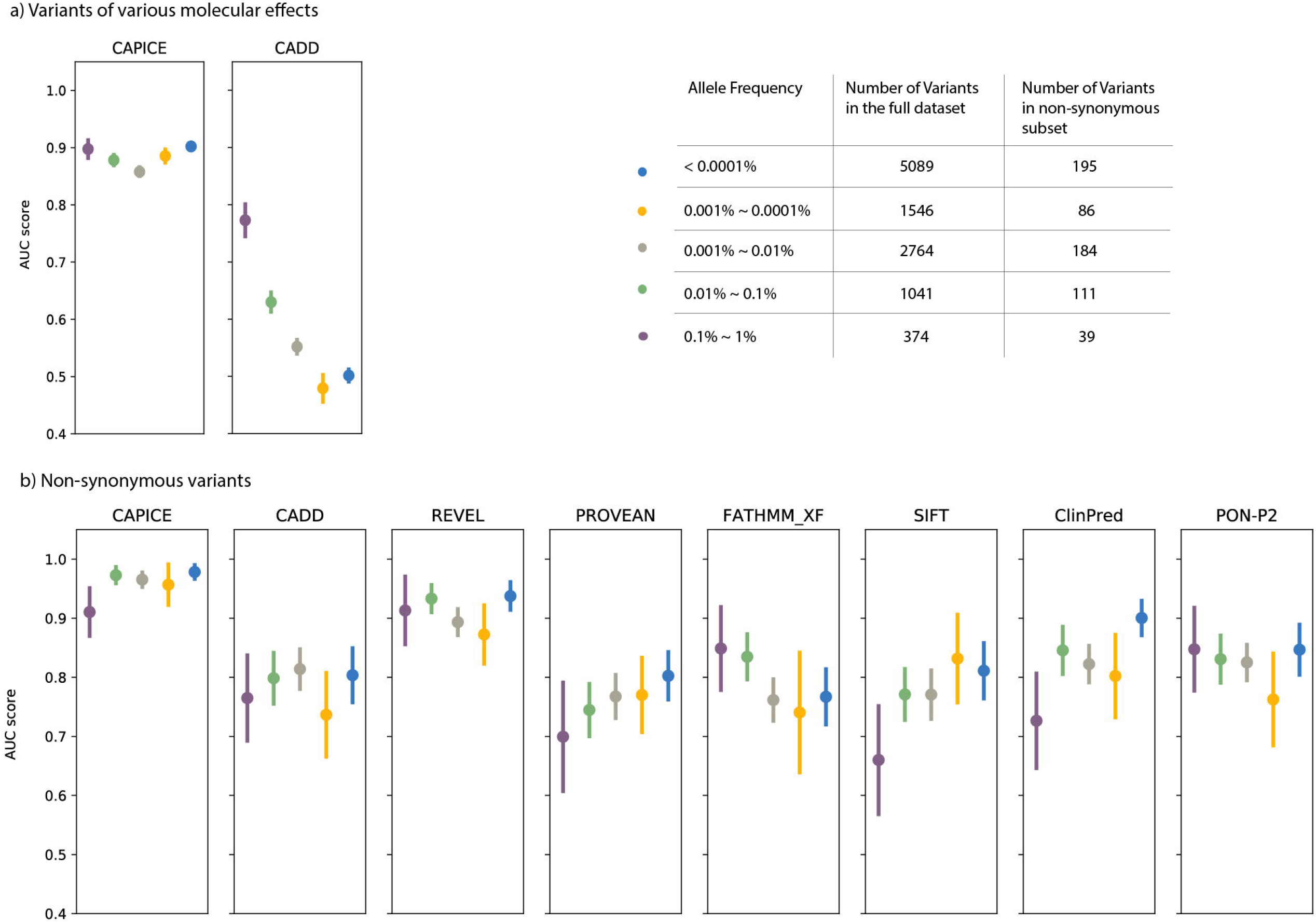
Performance comparison for rare and ultra-rare variants (a) for variants of different molecular consequences (b) in the missense subset. Each dot represents the mean AUC value with standard deviation.

For the full benchmark dataset, CAPICE performed consistently above 0.85 of AUC for variants with an allele frequency <1%, while the performance of CADD version 1.4 (30), the current best method for indicating the pathogenicity of variants throughout the genome compared to LINSIGHT (31), EIGEN (32), DeepSEA (33) drops significantly in case of rare variants (Figure 2*a*). For the non-synonymous subset, CAPICE consistently performed better than or comparably to the next-best method, REVEL, for variants within different allele frequency ranges, and better than all other methods (Figure *2b*).

For common variants (defined here as having an allele frequency >1%), the number of available pathogenic variants was too small (14 pathogenic variants) to get an accurate and robust performance measurement.

### CAPICE shows consistent prediction performance for different types of variants

CAPICE outperforms the current best computational prediction methods for variants that cause different molecular consequences (Figure *3* and Supplementary Figure *2*). For variants displaying different molecular consequences, CAPICE has an AUC of 0.92 for canonical splicing variants and an AUC of 0.97 for non-synonymous variants in the independent test dataset. Compared to CADD, CAPICE performs significantly better for multiple types of variants, particularly canonical splicing, stop-gained and frame-shift variants.

**Figure 3.**
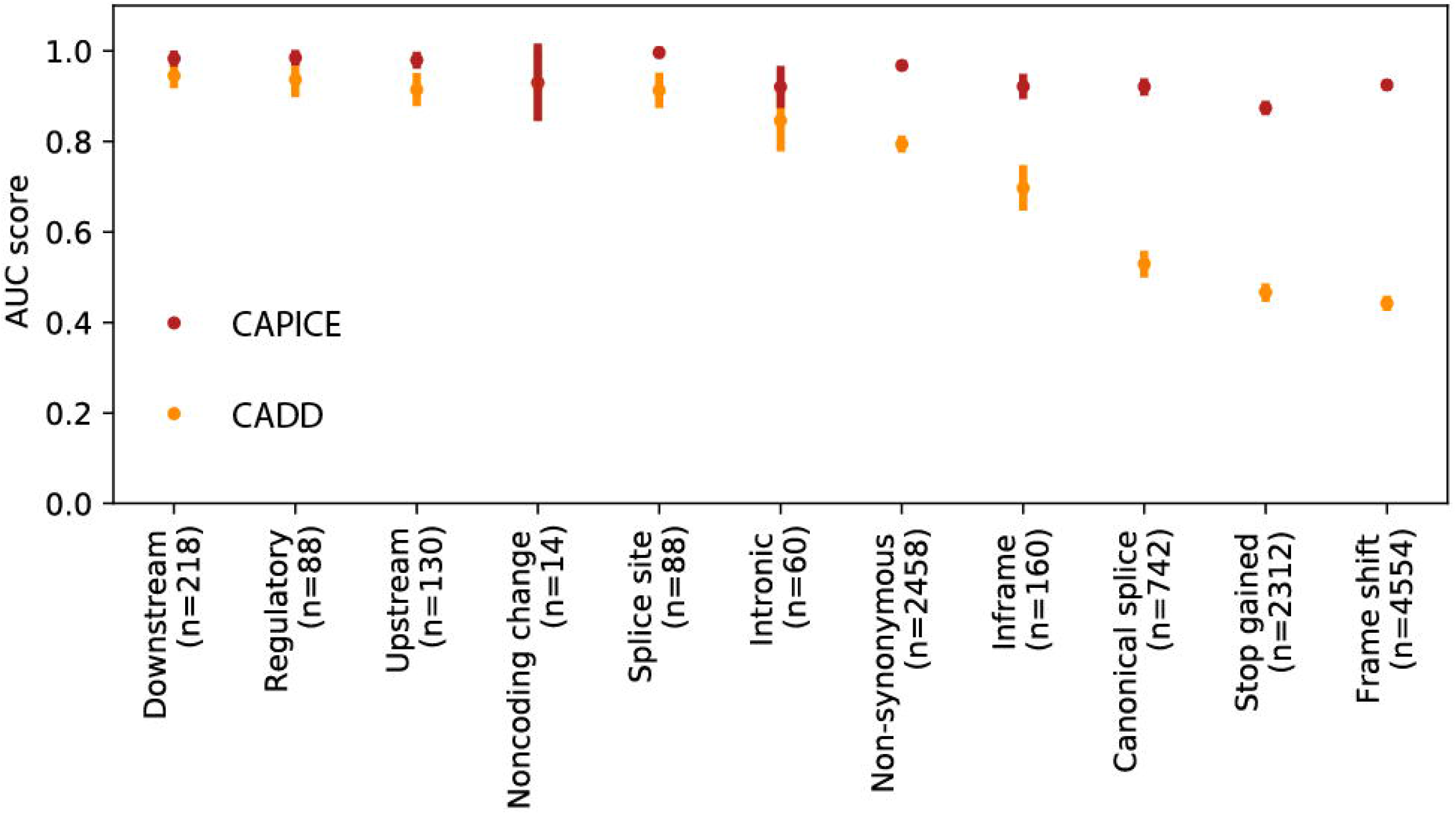
Performance comparison for variants of different molecular consequences of CAPICE and CADD.

**Figure 4.**
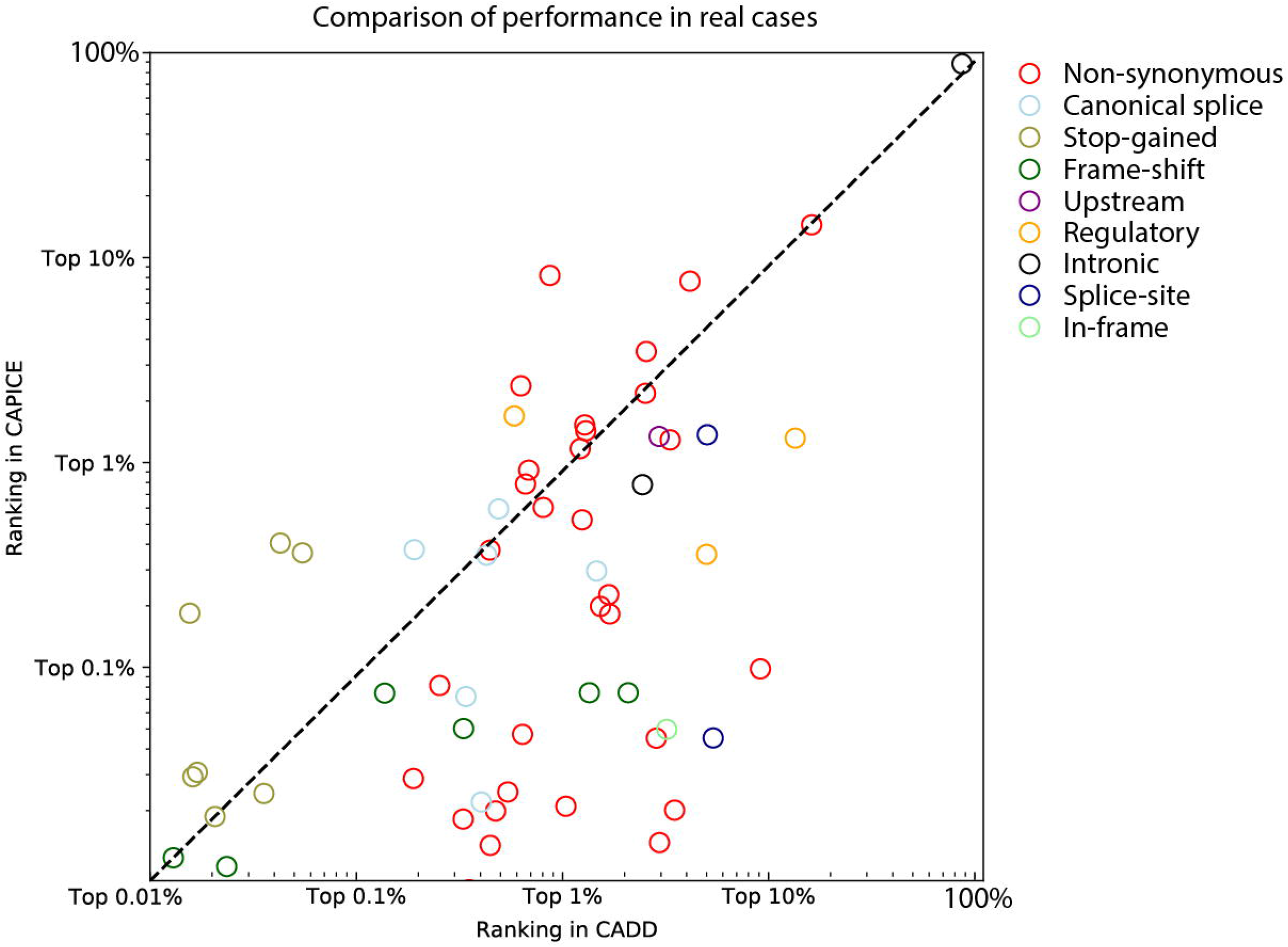
Performance comparison in real cases. In total, 54 patients and 58 variants were included. Each variant is reported as the diagnosis for that patient. Each dot in the plot shows a variant. The color of the dot represents the molecular effect predicted by VEP.

### CAPICE performance in clinical setting

In addition to the synthetic benchmark datasets, we also evaluated CAPICE’s performance in patients’ data.

To have first assessment of clinical utility, we used whole exome sequencing data from 54 solved patients from our diagnostics department and compared the ranking of the disease-causing variant with scores from CADD and CAPICE. We did not compare to REVEL, the second best method from our previous evaluation because a specific method for non-synonymous variants can miss variants of other molecular effects. A description of the solved patients’ can be found in (34). For each disease-causing variant discovered in that patient, we compared the performance of CAPICE and CADD by comparing the ranking of the particular variant among all variants observed within that patient. For 83% of the cases, CAPICE can prioritize the disease-causing variant within the 1% of the total variants observed in whole exome sequencing experiment, while CADD achieves the 1% performance for only 60% of the cases. Consistent with results described in previous sections that CAPICE achieves better AUC value for frameshift variants, CAPICE performed better for all cases with a disease-causing variant of frameshift effect.

## DISCUSSION

We have implemented a supervised machine learning approach called CAPICE to prioritize pathogenic SNVs and InDels for genomic diagnostics. CAPICE overcomes the limitations of existing methods, which either give predictions for a particular type of variants or showing moderate performance because they’re built for general purposes. We showed in multiple benchmark datasets, either derived from public databases or real patient cases that CAPICE outperforms the current best method for rare and ultra-rare variants with various molecular effects.

In this study, we used the same set of features as CADD used for constructing their score but trained the model directly on pathogenicity. The features enabled CAPICE to make predictions for variants of various molecular effects. Its focus on pathogenicity helped CAPICE to overcome the challenges faced by CADD in predicting pathogenicity (35) in the clinic. As a result, CAPICE gives significantly better prediction for rare variants, and various types of variants, in particular, frameshift, splicing, and stop-gained variants. We also observed that most current predictors have problems classifying rare and ultra-rare variants, with the exception for REVEL, an ensemble method that targets rare variants. We thus adopted the same strategy as REVEL by including rare variants when training CAPICE, and thereby obtained a comparable performance to that of REVEL for missense rare variants and significantly better results than all the other methods tested for ultra-rare variants.

We made full use of the large amount of data generated by other researchers. The evidence for a variant’s clinical relevance reported in public databases such as ClinVar can be conflicting or outdated (36). The star system used in ClinVar review status (37) serves as a good quality check for estimating the trustworthiness of the reported pathogenicity, and this quality estimation is used by many researchers as a selection criteria for constructing or evaluating variant prioritization methods [(15), (38)]. However, this method of data selection can introduce biases and waste potentially important information. In particular, neutral variants can be enriched for common ones. These common variants can be easily filtered out in a diagnostic pipeline using a general cut-off or expected carrier prevalence for specific diseases (39). Using such a biased dataset could however lead to a biased model or an overly optimistic performance estimation. When training CAPICE, we did not exclude lower-quality data, and assigned it a lower sample weight during model training. This strategy overcome the data selection bias mentioned above and led to a model with equally good performance for rare and ultra-rare variants. When testing CAPICE, we only selected high-quality data for the pathogenic set. For the neutral set, we included rare and ultra-rare variants for all the types of variations found in general population studies (after filtering for known pathogenic variations and inheritance mode). This allowed us to avoid the bias discussed above.

Current variant prioritization methods, including ours, often neglect context information about a patient such as phenotype information, family history and the cell types associated with specific diseases. Moreover, the methods developed are often evaluated in a stand-alone manner, and their associations with other steps in a genome diagnostic pipeline are not often investigated. In this study, we have only shown preliminary evaluation results using solved patient data. In future studies, we hope to include context information to further improve CAPICE’s predictive power. We also believe that the model’s performance needs to be discussed in a broader context that includes gene prioritization and mutational burden-testing.

## CONCLUSION

We have developed CAPICE, an ensemble method for prioritizing pathogenic variants in clinical exomes for Mendelian disorders, including SNVs and InDels, that outperforms all other existing methods and that we dream will greatly benefit rare disease research and patients worldwide. By re-using the CADD features, but training a machine-learning model on variants’ pathogenicity, CAPICE consistently outperforms other methods in our benchmark datasets for variants of various molecular effect and allele frequency. Additionally, we demonstrate that predictions made using CAPICE scores produce many fewer false positives than predictions made based on CADD scores. To enable its integration into automated and manual diagnostic pipelines, CAPICE is available as a free and open source software command-line tool from https://github.com/molgenis/capice and as a web-app at https://molgenis43.gcc.rug.nl/. Pre-computed scores are available as a download at https://doi.org/10.5281/zenodo.3516248.

## MATERIALS AND METHODS

The flowchart of this study is in Supplementary Figure *1*.

## DATA

### Data collection and selection

Training and benchmark data on neutral and pathogenic variants were derived from vcf files from the ClinVar database (17), dated 02 January 2019; from the VKGL data share consortium (40); from the GoNL data (41) and from data used in a previous study (29). From the ClinVar dataset, we collected variants reported by one or more submitters to have clear clinical significance, including pathogenic and likely pathogenic variants and neutral and likely neutral variants. From the VKGL data consortium, we collected variants with clear classifications, either (Likely) Pathogenic or (Likely) Benign, with support from one or more laboratories. The neutral variants from previous research developing the GAVIN tool (29) were mainly collected from ExAC without posing a constraint on allele frequency. We also obtained a neutral benchmark dataset from a benchmark study by (24).

In our data selection step, we removed duplicate variants located in unique chromosomal positions and those with inconsistent pathogenicity classification across the different databases. To reduce potential variants in general population datasets from carriers, we excluded variants observed in dominant genes using inheritance modes of each gene retrieved from the Clinical Genome Database dated 28 Feburary, 2019 (42).

In total, we collected 80k pathogenic variants and 450k putative neutral variants, and the entire dataset can be found in the Supplementary Material. After the initial cleaning step described above, we built a training dataset for model construction and a benchmark dataset that we left out of the training procedures so it could be used for performance evaluation later on.

### Construction of the benchmark and training sets

To build a benchmark dataset for performance evaluation that was fully independent of model construction procedures, we first selected the high-confidence pathogenic variants from the ClinVar and VKGL database. High-confidence variants are those with a review status of “two or more submitters providing assertion criteria provided the same interpretation (criteria provided, multiple submitters, no conflicts)”, “review by expert panel” and “practice guideline” in ClinVar database, and those are reported by one of more laboratories without conflicting interpretation in VKGL database. From the pathogenic variants that passed these criteria, we then randomly selected 50% to add into the benchmark dataset, which resulted in 6,937 pathogenic variants. To enable unbiased comparison of neutral and pathogenic variants with different molecular consequences, we created benchmark datasets with equal proportions of pathogenic and neutral variants for each type of molecular consequences, with the additional requirement that the pathogenic and neutral variants share similar distributions in allele frequency. An overview of the allele frequency distribution of the pathogenic and neutral variants for each type of molecular effects is in Supplementary Figure *2*.

In total, our benchmark set contained 10,842 variants and our training set contained 334,601 variants. The training set had 32,783 high confidence variants and 301,819 lower confidence variants. The high-confidence training variants were 12,646 pathogenic variants and 20,137 neutral variants. The lower confidence variants were 28,035 pathogenic variants and 273,784 neutral variants.

The two neutral benchmark datasets are those taken from a previous benchmark study and the GoNL dataset. The previous benchmark study by [(24)] selected neutral variants from the ExAC dataset and only included common variants with allele frequencies between 1% and 25%. For this dataset, we removed variants seen in the training set. In total, there were 60,699 neutral variants in our benchmark dataset. To build the neutral benchmark dataset from GoNL data, we selected all the variants that passed our quality assessment, then calculated their allele frequency within the GoNL population. We then selected those variants with an allele frequency <1% and removed variants that had been included in the training set. In total, there were 14,426,914 variants involved (Supplementary Table *2*).

### Data annotation and preprocessing

The collected variants in both the training and test datasets were annotated using CADD web service v1.4, which consists of 92 different features from VEP (version 90.5) (43) and epigenetic information from ENCODE (44) and the NIH RoadMap project (45). A detailed explanation of these features can be found in the (21) CADD paper. For each of the 11 categorical features, we selected up to five top levels to avoid introducing excessive sparsity, which could be computationally expensive, and used one-hot encoding before feeding the data into the model training procedures (46). For the 81 numerical variables, we imputed each feature using the imputation value recommended by (21). The allele frequency in the population was annotated using the vcfTool (47) from GnomAD r2.0.1 (48). We assigned variants not found in the GnomAD database an allele frequency of 0.

## MODEL CONSTRUCTION

### Model construction and training procedures

We trained a gradient-boosting tree model using the XGBoost (version 0.72) Python package. The hyper-parameters, n_estimators, max_depth and learning_rate were selected by 5-fold cross-validation using the RandomSearchCV function provided by the scikit-learn (version 0.19.1) Python package. Within each training fold, we used an early stopping criteria of 15 iterations. We then used the model trained with the best set of hyper-parameters (0.1 for learning_rate, 15 for max_depth and 422 for n_estimators) for performance measurement. For fitting the model, we also used the sample weight assigned to each variant. The sample weight is a score ranging from 0 to 1 that reflects the confidence level of the trustworthiness of the pathogenicity status of that variant. High-confidence variant, as described previously, are given a sample weight of 1, and the low-confidence variants were given a lower sample weight of 0.8. A variant with a high sample weight will thus contribute more to the loss function used in the training procedure (46). To test the assigned sample weights, we used the best set of parameters returned from the previous fine-tuning process and tried three different conditions in which we set the sample weights of the lower confidence variants to 0, 0.8 and 1. We then selected the model with the highest AUC value for the cross-validation dataset.

### Threshold-selection Strategies

For comparing the false positive rate in the neutral benchmark dataset and comparing the classification results, we tested different threshold-selection strategies for both CAPICE and CADD. For CAPICE, we obtained the threshold from the training dataset that results in a recall value within 0.94-0.96. To calculate the threshold, we searched for all possible threshold value from 0 to 1 and selected the first threshold for which the resulting recall value fall between 0.94 and 0.96. This method resulted in a general threshold of 0.02. For CADD, we tested two different threshold-selection methods. The first threshold was a default value of 20. The second method used GAVIN (29) to provide gene-specific thresholds. For other machine learning methods that returned a pathogenicity score ranging from 0 to 1, and no recommended threshold was given in the original paper, we selected a default value of 0.5. This includes the following methods: REVEL, ClinPred, SIFT and FATHMM-XF. For PROVEAN, we used a default score of -2.5 as the threshold.

## EVALUATION METRICS

For model performance comparison, we used Receiver Operating Characteristic (ROC) curve, AUC value (49), and measurements in the confusion matrix together with the threshold-selection strategies mentioned above. For measuring model performance in the neutral benchmark dataset, we examined the false positive rate. The false positive rate is the number of true neutral variants but predicted as pathogenic divided by the number of true neutral variants. To evaluate the robustness of the model predictions, we performed bootstrap on the benchmark dataset for standard deviation measurement for 100 repetitions, with the same sample size of the benchmark dataset for each repetition (50).

For evaluating performance in solved patients, we used the previously diagnosed patients with clear record of the disease-causing variant from University Medical Center in Groningen. A description of the solved patients’ can be found in (34). For examining CAPICEs performance, we first eliminated all variants with an allele frequency above 10% and then predicted the pathogenicity for the remaining variants. Subsequently, we sorted the variants of each individual by their pathogenicity score assigned by the respective predictors, and used the ranking of the disease-causing variant found within that individual as the measurement.

The data and pathogenicity predictions are provided in Web resources.

## Data Availability

CAPICE is available as a free and open source software command-line tool from https://github.com/molgenis/capice and as a web-app at https://molgenis43.gcc.rug.nl/. Pre-computed scores are available as a download at https://doi.org/10.5281/zenodo.3516248

## WEB RESOURCE

CAPICE’s Precomputed Scores: https://doi.org/10.5281/zenodo.3516248

CAPICE: https://github.com/molgenis/capice and web application https://molgenis43.gcc.rug.nl

CADD: https://cadd.gs.washington.edu/score

REVEL: https://sites.google.com/site/revelgenomics/

PON-P2: http://structure.bmc.lu.se/PON-P2/

ClinPred: https://sites.google.com/site/clinpred/

PROVEAN and SIFT: http://provean.jcvi.org/genome_submit_2.php?species=human

GAVIN: https://molgenis.org/gavin

FATHMM-XF: http://fathmm.biocompute.org.uk/fathmm-xf/

## DECLARATIONS

All manuscripts must contain the following sections under the heading ‘Declarations’:

### Ethics approval and consent to participate

Not Applicable

### Consent for publication

Not Applicable

### Availability of data and materials

Training and testing data with label and predictions from CAPICE and tested predictors and the pre-computed scores for all possible SNVs and InDels is available online at Zenodo: https://doi.org/10.5281/zenodo.3516248 and at Github: https://github.com/molgenis/capice.

### Authors’ contributions

Shuang Li, K. Joeri van der Velde, Morris A. Swertz designed the experiments, analyzed the data, and wrote the paper. K, Joeri van der Velde and Morris A. Swertz provided support in supervising Shuang Li in conducting the projects. Dick de Ridder, Aalt-Jan van Dijk, Dimitrios Soudis, and Leslie Zwerwer provided support for experimental design and model construction. Patrick Deelen provided support for experiment design and evaluation of the model in real patients’ data. Dennis Hendriksen and Bart Charbon provided support for web application and web service API construction. Marielle van Gijn and Richard Sinke provided support for interpreting the results. Kristin Abbot, Birgit Sikkema, Cleo van Diemen, Mieke Kerstjens-Frederikse provided support in collecting the patients’ diagnostic records and interpreting the results. All authors read and approved the final manuscript.

## Acknowledgements

Kate Mc Intyre contributed greatly in the development and refinement of texts. Harm-Jan Westra helped us in reviewing the manuscript. Tommy de Boer provided support with the web service API construction.

## SUPPLEMENTARY MATERIALS

### Additional Figures and Tables

**Supplementary Figure 1.**
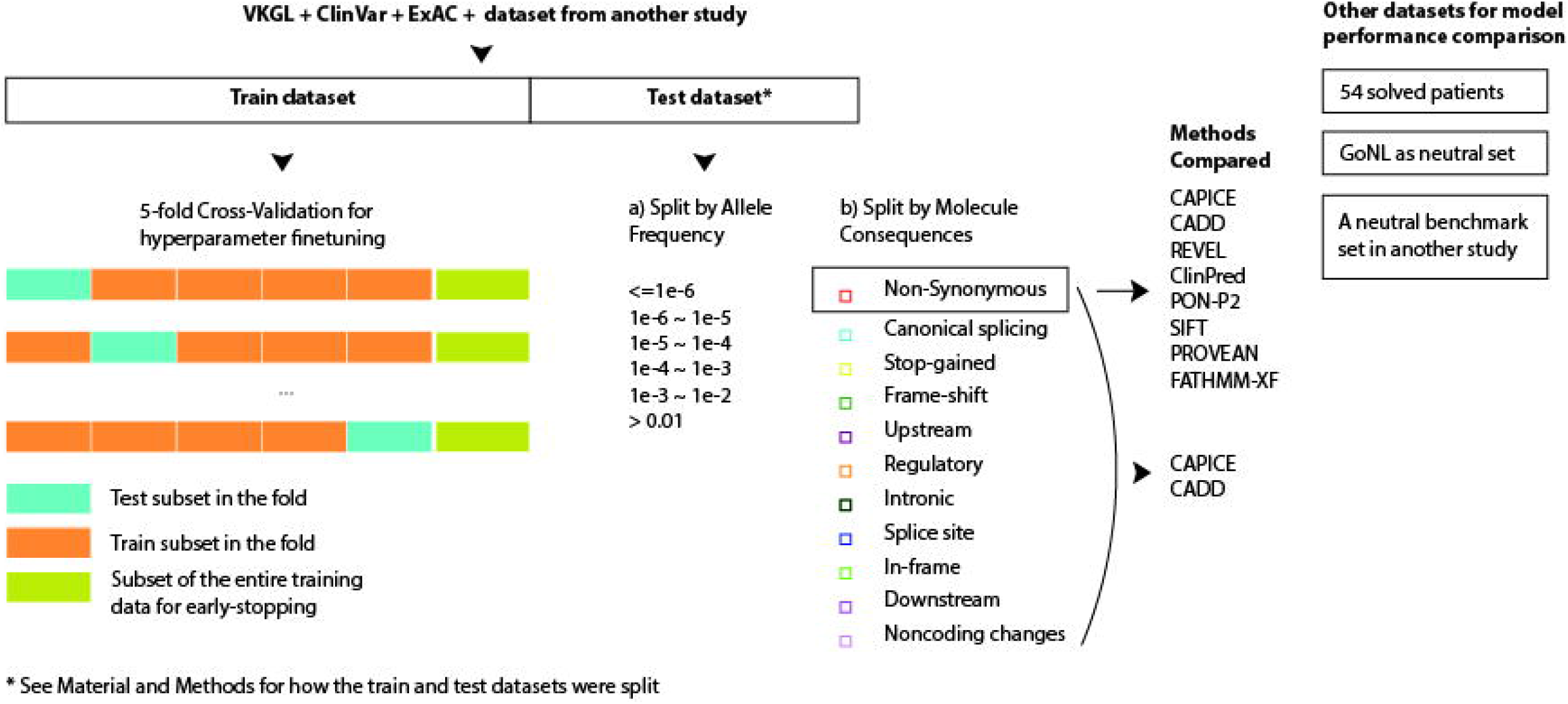
Flowchart of this study

**Supplementary Figure 2.**
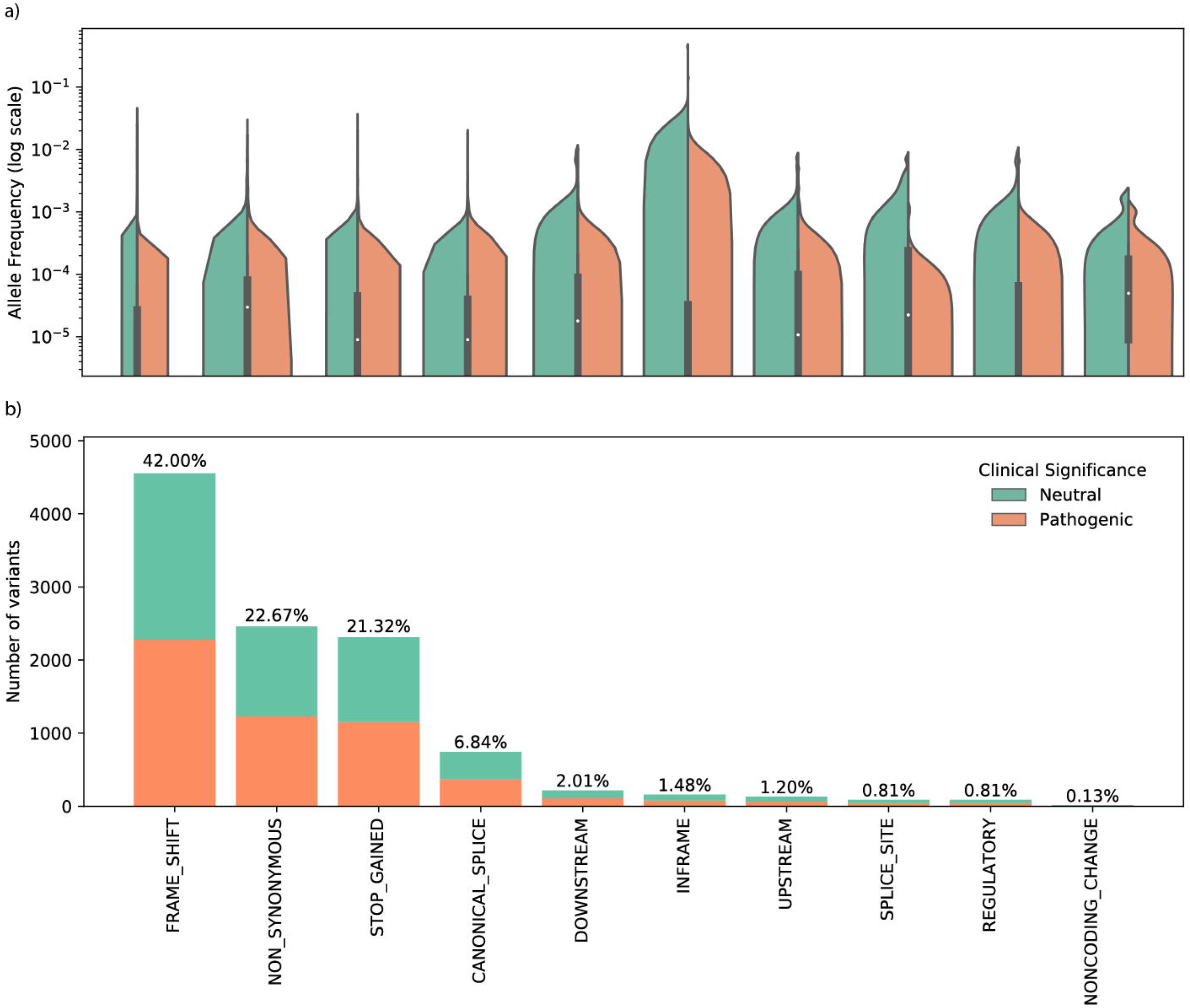
For benchmarking, we created a balanced test dataset that is equally distributed in terms of a) the number of pathogenic and putatively neutral variants and b) allele frequency distribution for different molecular consequences.

**Supplementary Figure 3.**
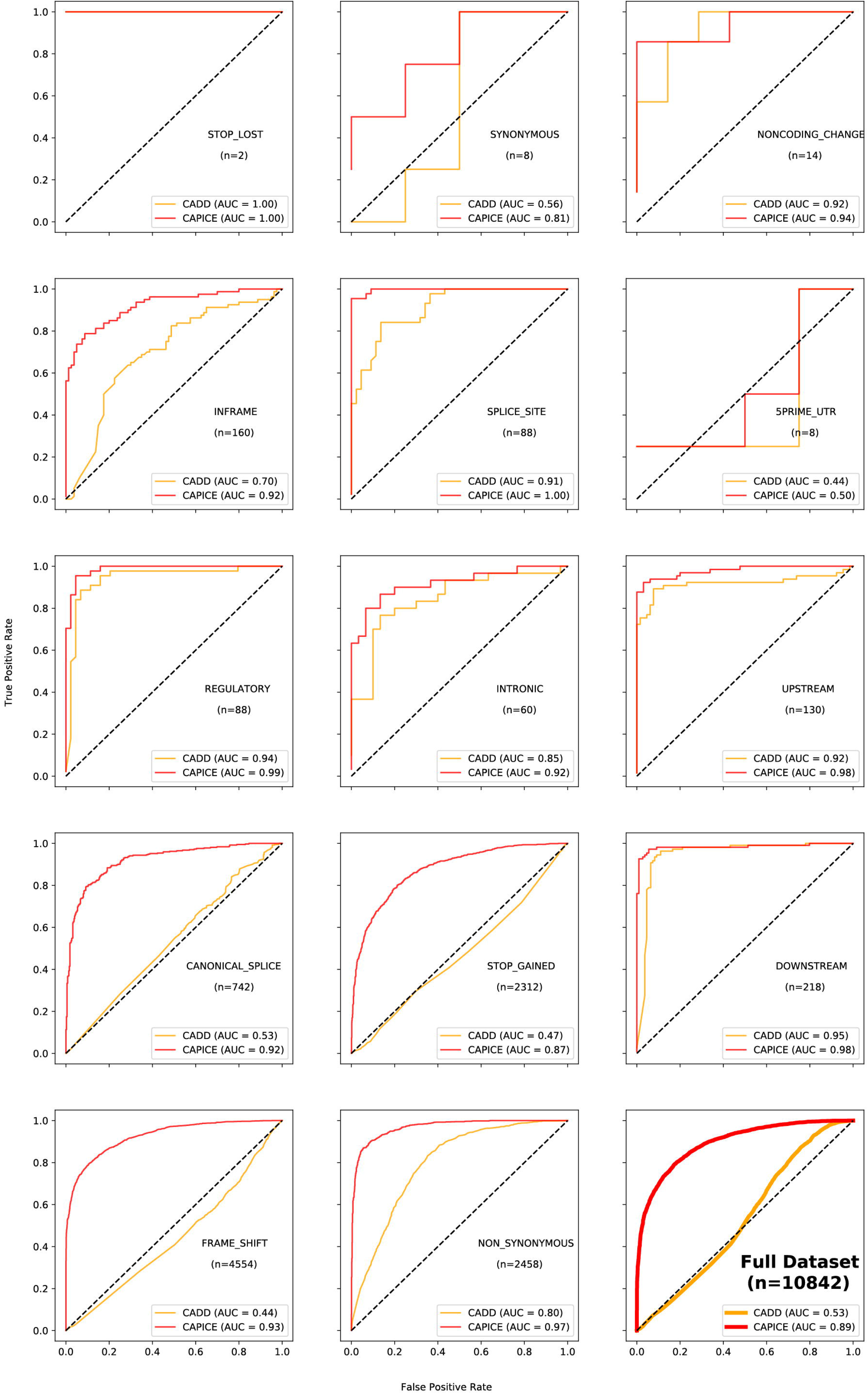
ROC curves and AUC values from CAPICE and CADD for variants with different molecular functions.

**Supplementary Figure 4.**
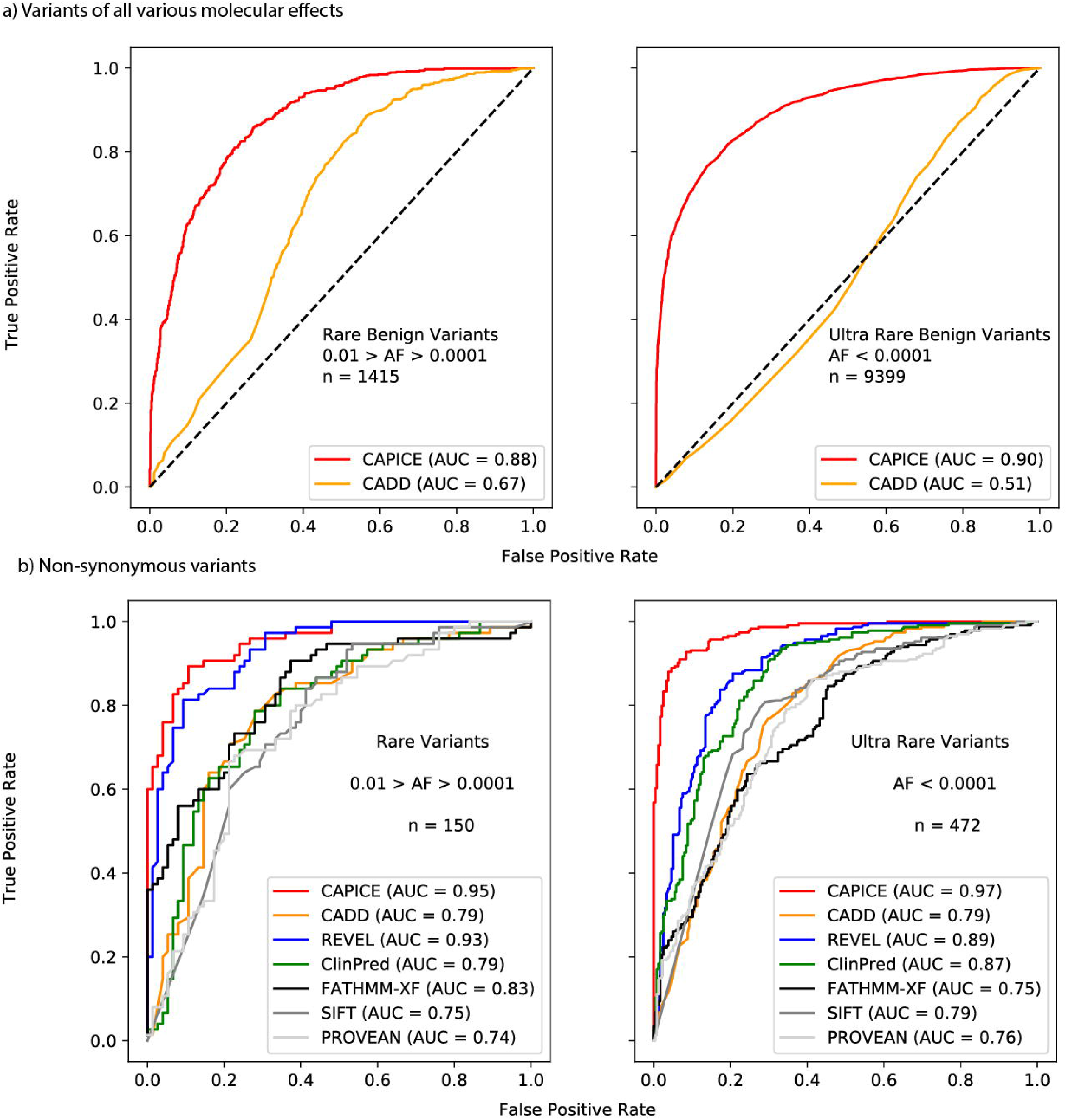
AUC and ROC curves for the a) full dataset and b) missense subset for rare and ultra-rare variants defined as variants with allele frequency between 0.01% and 0.1% and variants with allele frequency <0.01%

**Supplementary Table 1.** Description of all methods tested in the study

| Method name | Application | Link for web resources |
| --- | --- | --- |
| CADD | Estimating the relative pathogenicity of SNVs and InDels | https://cadd.gs.washington.edu/ |
| REVEL | Predicting the pathogenicity of rare missense variants | https://sites.google.com/site/revelgenomics/ |
| ClinPred | Predicting the pathogenicity of missense variants | https://sites.google.com/site/clinpred/ |
| PON-P2 | Predicting the pathogenicity of missense variants | http://structure.bmc.lu.se/PON-P2/ |
| SIFT | Predicting missense variants effects on protein function | http://provean.jcvi.org/genome_submit_2.php?species=human |
| PROVEAN | Predicting missense variants and InDels' effects on protein function | http://provean.jcvi.org/genome_submit_2.php?species=human |
| FATHMM-XF | Predicting pathogenicity of point mutations | http://fathmm.biocompute.org.uk/fathmm-xf/ |

**Supplementary Table 2.** CAPICE and CADD false positive rates in the neutral benchmark dataset

| Molecular Consequence | CADD (20) | CIPICE (General Threshold) | Number of Variants |
| --- | --- | --- | --- |
| NON_SYNONYMOUS | 0.37 | 0.07 | 50181 |
| DOWNSTREAM | 0.36 | 0.11 | 12121 |
| REGULATORY | 0.35 | 0.12 | 11317 |
| UPSTREAM | 0.35 | 0.1 | 10959 |
| INTRONIC | 0.25 | 0.06 | 10910 |
| NONCODING_CHANGE | 0.34 | 0.06 | 1202 |
| 3PRIME_UTR | 0.23 | 0.03 | 902 |
| SYNONYMOUS | 0.07 | 0.08 | 456 |
| 5PRIME_UTR | 0.19 | 0.03 | 295 |
| SPLICE_SITE | 0.15 | 0.08 | 137 |
| CANONICAL_SPLICE | 0.6 | 0.4 | 10 |
| STOP_GAINED | 1 | 0.22 | 9 |

**Supplementary Table 3.** CAPICE and CADD false positive rates in the GoNL dataset

|  | CADD (20) | CIPICE | Number of Variants |
| --- | --- | --- | --- |
| INTRONIC | 0.00 | 0.00 | 6483483 |
| INTERGENIC | 0.00 | 0.00 | 4399474 |
| DOWNSTREAM | 0.01 | 0.00 | 1205398 |
| UPSTREAM | 0.01 | 0.00 | 1174342 |
| REGULATORY | 0.01 | 0.01 | 808360 |
| NONCODING_CHANGE | 0.01 | 0.00 | 115099 |
| 3PRIME_UTR | 0.02 | 0.00 | 108194 |
| NON_SYNONYMOUS | 0.57 | 0.19 | 67013 |
| SYNONYMOUS | 0.02 | 0.08 | 36177 |
| 5PRIME_UTR | 0.03 | 0.01 | 14526 |
| SPLICE_SITE | 0.05 | 0.08 | 10352 |
| CANONICAL_SPLICE | 0.58 | 0.54 | 1545 |
| STOP_GAINED | 1.00 | 0.74 | 1647 |
| FRAME_SHIFT | 0.88 | 0.9 | 821 |
| INFRAME | 0.37 | 0.63 | 415 |
| STOP_LOST | 0.06 | 0.37 | 68 |

## REFERENCES

1. Boudellioua I, Mahamad Razali RB, Kulmanov M, Hashish Y, Bajic VB, Goncalves-Serra E, et al. Semantic prioritization of novel causative genomic variants. PLoS Comput Biol [Internet]. 2017 Apr [cited 2018 May 3];13(4):e1005500. Available from: http://www.ncbi.nlm.nih.gov/pubmed/28414800

2. Lionel AC, Costain G, Monfared N, Walker S, Reuter MS, Hosseini SM, et al. Improved diagnostic yield compared with targeted gene sequencing panels suggests a role for whole-genome sequencing as a first-tier genetic test. Genet Med [Internet]. 2018 Apr 3 [cited 2018 May 9];20(4):435–43. Available from: http://www.nature.com/doifinder/10.1038/gim.2017.119

3. Clark MM, Hildreth A, Batalov S, Ding Y, Chowdhury S, Watkins K, et al. Diagnosis of genetic diseases in seriously ill children by rapid whole-genome sequencing and automated phenotyping and interpretation. Sci Transl Med [Internet]. 2019 Apr 24 [cited 2019 Oct 2];11(489):eaat6177. Available from: http://www.ncbi.nlm.nih.gov/pubmed/31019026

4. Sawyer SL, Hartley T, Dyment DA, Beaulieu CL, Schwartzentruber J, Smith A, et al. Utility of whole-exome sequencing for those near the end of the diagnostic odyssey: time to address gaps in care. Clin Genet [Internet]. 2016 Mar [cited 2019 Oct 2];89(3):275–84. Available from: http://www.ncbi.nlm.nih.gov/pubmed/26283276

5. Trujillano D, Bertoli-Avella AM, Kumar Kandaswamy K, Weiss ME, Köster J, Marais A, et al. Clinical exome sequencing: results from 2819 samples reflecting 1000 families. Eur J Hum Genet [Internet]. 2017 Feb 16 [cited 2018 Nov 30];25(2):176–82. Available from: http://www.nature.com/articles/ejhg2016146

6. Meng L, Pammi M, Saronwala A, Magoulas P, Ghazi AR, Vetrini F, et al. Use of Exome Sequencing for Infants in Intensive Care Units. JAMA Pediatr [Internet]. 2017 Dec 4 [cited 2019 Oct 2];171(12):e173438. Available from: http://www.ncbi.nlm.nih.gov/pubmed/28973083

7. Bardakjian TM, Helbig I, Quinn C, Elman LB, Mccluskey LF, Scherer SS, et al. Genetic test utilization and diagnostic yield in adult patients with neurological disorders. [cited 2018 Nov 30]; Available from: https://doi.org/10.1007/s10048-018-0544-x

8. Eilbeck K, Quinlan A, Yandell M. Settling the score: variant prioritization and Mendelian disease. Nat Rev Genet [Internet]. 2017 Aug 14 [cited 2018 Jan 31];18(10):599–612. Available from: http://www.nature.com/doifinder/10.1038/nrg.2017.52

9. Thiffault I, Farrow E, Zellmer L, Berrios C, Miller N, Gibson M, et al. Clinical genome sequencing in an unbiased pediatric cohort. Genet Med [Internet]. 2019 Feb 16 [cited 2019 Oct 2];21(2):303–10. Available from: http://www.nature.com/articles/s41436-018-0075-8

10. Berberich AJ, Ho R, Hegele RA. Whole genome sequencing in the clinic: empowerment or too much information? CMAJ [Internet]. 2018 [cited 2019 Oct 2];190(5):E124–5. Available from: http://www.ncbi.nlm.nih.gov/pubmed/29431109

11. Shi F, Yao Y, Bin Y, Zheng C-H, Xia J. Computational identification of deleterious synonymous variants in human genomes using a feature-based approach. BMC Med Genomics [Internet]. 2019 Jan 31 [cited 2019 Oct 2];12(S1):12. Available from: https://bmcmedgenomics.biomedcentral.com/articles/10.1186/s12920-018-0455-6

12. Jagadeesh KA, Paggi JM, Ye JS, Stenson PD, Cooper DN, Bernstein JA, et al. S-CAP extends pathogenicity prediction to genetic variants that affect RNA splicing. Nat Genet [Internet]. 2019 Apr 25 [cited 2019 Oct 2];51(4):755–63. Available from: http://www.nature.com/articles/s41588-019-0348-4

13. Rogers MF, Shihab HA, Mort M, Cooper DN, Gaunt TR, Campbell C. FATHMM-XF: accurate prediction of pathogenic point mutations via extended features. Hancock J, editor. Bioinformatics [Internet]. 2018 Feb 1 [cited 2019 Oct 2];34(3):511–3. Available from: http://www.ncbi.nlm.nih.gov/pubmed/28968714

14. Ng PC, Henikoff S. SIFT: Predicting amino acid changes that affect protein function. Nucleic Acids Res [Internet]. 2003 Jul 1 [cited 2019 Oct 2];31(13):3812–4. Available from: http://www.ncbi.nlm.nih.gov/pubmed/12824425

15. Alirezaie N, Kernohan KD, Hartley T, Majewski J, Hocking TD. ClinPred: Prediction Tool to Identify Disease-Relevant Nonsynonymous Single-Nucleotide Variants. Am J Hum Genet [Internet]. 2018 Oct 4 [cited 2019 Oct 2];103(4):474–83. Available from: http://www.ncbi.nlm.nih.gov/pubmed/30220433

16. Ioannidis NM, Rothstein JH, Pejaver V, Middha S, McDonnell SK, Baheti S, et al. REVEL: An Ensemble Method for Predicting the Pathogenicity of Rare Missense Variants. Am J Hum Genet [Internet]. 2016 Oct 6 [cited 2019 Oct 2];99(4):877–85. Available from: http://www.ncbi.nlm.nih.gov/pubmed/27666373

17. Landrum MJ, Lee JM, Riley GR, Jang W, Rubinstein WS, Church DM, et al. ClinVar: public archive of relationships among sequence variation and human phenotype. Nucleic Acids Res [Internet]. 2014 Jan [cited 2019 Oct 2];42(Database issue):D980-5. Available from: http://www.ncbi.nlm.nih.gov/pubmed/24234437

18. Siepel A, Bejerano G, Pedersen JS, Hinrichs AS, Hou M, Rosenbloom K, et al. Evolutionarily conserved elements in vertebrate, insect, worm, and yeast genomes. Genome Res [Internet]. 2005 Aug 1 [cited 2019 Oct 2];15(8):1034–50. Available from: http://www.ncbi.nlm.nih.gov/pubmed/16024819

19. Davydov E V., Goode DL, Sirota M, Cooper GM, Sidow A, Batzoglou S. Identifying a High Fraction of the Human Genome to be under Selective Constraint Using GERP++. Wasserman WW, editor. PLoS Comput Biol [Internet]. 2010 Dec 2 [cited 2019 Oct 2];6(12):e1001025. Available from: https://dx.plos.org/10.1371/journal.pcbi.1001025

20. Quang D, Chen Y, Xie X. DANN: a deep learning approach for annotating the pathogenicity of genetic variants. Bioinformatics [Internet]. 2015 Mar 1 [cited 2019 Oct 2];31(5):761–3. Available from: https://academic.oup.com/bioinformatics/article-lookup/doi/10.1093/bioinformatics/btu703

21. Kircher M, Witten DM, Jain P, O’Roak BJ, Cooper GM, Shendure J. A general framework for estimating the relative pathogenicity of human genetic variants. Nat Genet [Internet]. 2014 Mar 2 [cited 2019 Oct 2];46(3):310–5. Available from: http://www.nature.com/articles/ng.2892

22. Rentzsch P, Witten D, Cooper GM, Shendure J, Kircher M. CADD: predicting the deleteriousness of variants throughout the human genome. Nucleic Acids Res [Internet]. 2019 Jan 8 [cited 2019 Oct 2];47(D1):D886–94. Available from: https://academic.oup.com/nar/article/47/D1/D886/5146191

23. Fu W, O’Connor TD, Jun G, Kang HM, Abecasis G, Leal SM, et al. Analysis of 6,515 exomes reveals the recent origin of most human protein-coding variants. Nature [Internet]. 2013 Jan 28 [cited 2019 Oct 2];493(7431):216–20. Available from: http://www.nature.com/articles/nature11690

24. Niroula A, Vihinen M. How good are pathogenicity predictors in detecting benign variants? Panchenko ARR, editor. PLOS Comput Biol [Internet]. 2019 Feb 11 [cited 2019 Oct 2];15(2):e1006481. Available from: http://dx.plos.org/10.1371/journal.pcbi.1006481

25. Ghosh R, Oak N, Plon SE. Evaluation of in silico algorithms for use with ACMG/AMP clinical variant interpretation guidelines. Genome Biol [Internet]. 2017 Dec 28 [cited 2018 Jan 15];18(1):225. Available from: https://genomebiology.biomedcentral.com/articles/10.1186/s13059-017-1353-5

26. Dong C, Wei P, Jian X, Gibbs R, Boerwinkle E, Wang K, et al. Comparison and integration of deleteriousness prediction methods for nonsynonymous SNVs in whole exome sequencing studies. Hum Mol Genet [Internet]. 2015 Apr 15 [cited 2018 May 7];24(8):2125–37. Available from: https://academic.oup.com/hmg/article-lookup/doi/10.1093/hmg/ddu733

27. Schaafsma GCP, Vihinen M. VariSNP, A Benchmark Database for Variations From dbSNP. Hum Mutat [Internet]. 2015 Feb [cited 2019 Oct 2];36(2):161–6. Available from: http://doi.wiley.com/10.1002/humu.22727

28. Sarkar A, Yang Y, Vihinen M. Variation Benchmark Datasets: Update, Criteria, Quality and Applications. bioRxiv [Internet]. 2019 May 10 [cited 2019 Oct 2];634766. Available from: https://www.biorxiv.org/content/10.1101/634766v1

29. van der Velde KJ, de Boer EN, van Diemen CC, Sikkema-Raddatz B, Abbott KM, Knopperts A, et al. GAVIN: Gene-Aware Variant INterpretation for medical sequencing. Genome Biol [Internet]. 2017 Dec 16 [cited 2019 Oct 2];18(1):6. Available from: http://genomebiology.biomedcentral.com/articles/10.1186/s13059-016-1141-7

30. Rentzsch P, Witten D, Cooper GM, Shendure J, Kircher M. CADD: predicting the deleteriousness of variants throughout the human genome. Nucleic Acids Res [Internet]. 2019 Jan 8 [cited 2019 Oct 2];47(D1):D886–94. Available from: http://www.ncbi.nlm.nih.gov/pubmed/30371827

31. Huang Y-F, Gulko B, Siepel A. Fast, scalable prediction of deleterious noncoding variants from functional and population genomic data. Nat Genet [Internet]. 2017 Mar 13 [cited 2018 Jan 15];49(4):618–24. Available from: http://www.nature.com/doifinder/10.1038/ng.3810

32. Ionita-Laza I, McCallum K, Xu B, Buxbaum JD. A spectral approach integrating functional genomic annotations for coding and noncoding variants. Nat Genet [Internet]. 2016 Feb [cited 2019 Oct 23];48(2):214–20. Available from: http://www.ncbi.nlm.nih.gov/pubmed/26727659

33. Zhou J, Troyanskaya OG. Predicting effects of noncoding variants with deep learning-based sequence model. Nat Methods. 2015 Sep 29;12(10):931–4.

34. Deelen P, van Dam S, Herkert JC, Karjalainen JM, Brugge H, Abbott KM, et al. Improving the diagnostic yield of exome-sequencing by predicting gene–phenotype associations using large-scale gene expression analysis. Nat Commun [Internet]. 2019 Dec 28 [cited 2019 Oct 2];10(1):2837. Available from: http://www.nature.com/articles/s41467-019-10649-4

35. Mather CA, Mooney SD, Salipante SJ, Scroggins S, Wu D, Pritchard CC, et al. CADD score has limited clinical validity for the identification of pathogenic variants in noncoding regions in a hereditary cancer panel. Genet Med [Internet]. 2016 Dec 5 [cited 2019 Oct 2];18(12):1269–75. Available from: http://www.nature.com/articles/gim201644

36. Shah N, Hou Y-CC, Yu H-C, Sainger R, Caskey CT, Venter JC, et al. Identification of Misclassified ClinVar Variants via Disease Population Prevalence. Am J Hum Genet [Internet]. 2018 Apr [cited 2019 Oct 2];102(4):609–19. Available from: https://linkinghub.elsevier.com/retrieve/pii/S0002929718300879

37. Review status in ClinVar [Internet]. [cited 2019 Oct 2]. Available from: https://www.ncbi.nlm.nih.gov/clinvar/docs/review_status/

38. Bao R, Huang L, Andrade J, Tan W, Kibbe WA, Jiang H, et al. Review of current methods, applications, and data management for the bioinformatics analysis of whole exome sequencing. Cancer Inform [Internet]. 2014 [cited 2018 Jan 19];13(Suppl 2):67–82. Available from: http://www.ncbi.nlm.nih.gov/pubmed/25288881

39. Richards S, Aziz N, Bale S, Bick D, Das S, Gastier-Foster J, et al. Standards and guidelines for the interpretation of sequence variants: a joint consensus recommendation of the American College of Medical Genetics and Genomics and the Association for Molecular Pathology. 2015 [cited 2018 Jan 15]; Available from: https://www.acmg.net/docs/Standards_Guidelines_for_the_Interpretation_of_Sequence_Variants.pdf

40. Fokkema IFAC, Velde KJ, Slofstra MK, Ruivenkamp CAL, Vogel MJ, Pfundt R, et al. Dutch genome diagnostic laboratories accelerated and improved variant interpretation and increased accuracy by sharing data. Hum Mutat [Internet]. 2019 Sep 3 [cited 2019 Oct 15];humu.23896. Available from: https://onlinelibrary.wiley.com/doi/abs/10.1002/humu.23896

41. Boomsma DI, Wijmenga C, Slagboom EP, Swertz MA, Karssen LC, Abdellaoui A, et al. The Genome of the Netherlands: design, and project goals. Eur J Hum Genet [Internet]. 2014 Feb [cited 2019 Oct 15];22(2):221–7. Available from: http://www.nature.com/articles/ejhg2013118

42. Solomon BD, Nguyen A-D, Bear KA, Wolfsberg TG. Clinical Genomic Database. Proc Natl Acad Sci [Internet]. 2013 Jun 11 [cited 2019 Oct 15];110(24):9851–5. Available from: http://www.pnas.org/cgi/doi/10.1073/pnas.1302575110

43. McLaren W, Gil L, Hunt SE, Riat HS, Ritchie GRS, Thormann A, et al. The Ensembl Variant Effect Predictor. Genome Biol [Internet]. 2016 Dec 6 [cited 2019 Oct 2];17(1):122. Available from: http://genomebiology.biomedcentral.com/articles/10.1186/s13059-016-0974-4

44. ENCODE Project Consortium TEP. An integrated encyclopedia of DNA elements in the human genome. Nature [Internet]. 2012 Sep 6 [cited 2019 Oct 2];489(7414):57–74. Available from: http://www.ncbi.nlm.nih.gov/pubmed/22955616

45. Bernstein BE, Stamatoyannopoulos JA, Costello JF, Ren B, Milosavljevic A, Meissner A, et al. The NIH Roadmap Epigenomics Mapping Consortium. Nat Biotechnol [Internet]. 2010 Oct [cited 2019 Oct 2];28(10):1045–8. Available from: http://www.ncbi.nlm.nih.gov/pubmed/20944595

46. Chen T, Guestrin C. XGBoost. In: Proceedings of the 22nd ACM SIGKDD International Conference on Knowledge Discovery and Data Mining - KDD ‘16 [Internet]. New York, New York, USA: ACM Press; 2016 [cited 2019 Oct 2]. p. 785–94. Available from: http://dl.acm.org/citation.cfm?doid=2939672.2939785

47. Danecek P, Auton A, Abecasis G, Albers CA, Banks E, DePristo MA, et al. The variant call format and VCFtools. Bioinformatics [Internet]. 2011 Aug 1 [cited 2019 Oct 2];27(15):2156–8. Available from: http://www.ncbi.nlm.nih.gov/pubmed/21653522

48. Karczewski KJ, Francioli LC, Tiao G, Cummings BB, Alföldi J, Wang Q, et al. Variation across 141,456 human exomes and genomes reveals the spectrum of loss-of-function intolerance across human protein-coding genes. bioRxiv [Internet]. 2019 [cited 2019 Oct 24];531210. Available from: https://www.biorxiv.org/content/10.1101/531210v2

49. Hanley JA, McNeil BJ. The meaning and use of the area under a receiver operating characteristic (ROC) curve. Radiology [Internet]. 1982 Apr [cited 2019 Oct 2];143(1):29–36. Available from: http://www.ncbi.nlm.nih.gov/pubmed/7063747

50. Bishop CM. Pattern Recognition And Machine Learning - Springer 2006. 2006.

